# Human lung tissue resident memory T cells are re-programmed but not eradicated with systemic glucocorticoids after acute cellular rejection

**DOI:** 10.1101/2021.09.27.21263985

**Authors:** Mark E. Snyder, Kaveh Moghbeli, Anna Bondonese, Andrew Craig, Iulia Popescu, Li Fan, Tracy Tabib, Robert Lafyatis, Kong Chen, Humberto E. Trejo Bittar, Elizabeth Lendermon, Joseph Pilewski, Bruce Johnson, Silpa Kilaru, Yingze Zhang, Pablo G. Sanchez, Jonathan K. Alder, Peter A. Sims, John F. McDyer

## Abstract

Acute cellular rejection is common after lung transplantation and is associated with an increased risk of early chronic rejection. We present combined single cell RNA and T cell receptor sequencing on recipient derived T cells obtained from the bronchoalveolar lavage of three lung transplant recipients with acute cellular rejection and compare them with T cells obtained from the same three patients after clinical treatment of rejection with high-dose, systemic glucocorticoids. At the time of acute cellular rejection, we find an oligoclonal expansion of cytotoxic CD8^+^ T cells, that all persist as tissue resident memory T cells following successful treatment. Persisting CD8^+^ allograft-resident T cells have reduced gene expression for cytotoxic mediators following therapy with glucocorticoids. This clonal expansion is discordant with circulating T cell clonal expansion at the time of rejection, suggesting in-situ expansion. These findings pose a potential biological mechanism linking acute cellular rejection to chronic allograft damage.

## Introduction

Acute cellular rejection (ACR) is common following lung transplantation, occurring in between 29% - 55% of recipients within the first year of transplantation^1,2^. In addition to contributing to patient morbidity, ACR is associated with an increased risk of early chronic lung allograft dysfunction (CLAD), a progressive fibrosis of the small airways and the major limiting factor to long-term survival following lung transplantation. ACR is defined in lung transplant recipients as grades of perivascular or peri-bronchial infiltrates, predominantly composed of lymphocytes, found at the time of transbronchial biopsy. T cells are the predominant mediator of ACR in solid organ transplantation including the lung^3,4^. Alloreactive T cells can be formed after priming by donor peptide presented by donor antigen presenting cells (referred to as the direct pathway), donor peptide presented by recipient antigen presenting cells (indirect pathway), or donor peptide presented on donor major histocompatibility complexes taken up by recipient antigen presenting cells (semi-direct pathway)^5–8^. The first-line treatment for ACR consists of high doses of glucocorticoids and/or augmentation of maintenance immunosuppression^9^. In cases of advanced grade ACR or ACR refractory to high dose systemic glucocorticoids, lympho-depletive therapies such as anti-thymocyte globulin or monoclonal antibody to CD52 (alemtuzumab) can be effective treatment options^10^.

Immediately following transplantation, circulating, recipient-derived T cells begin to populate the lung allograft while the proportion of donor-derived tissue resident memory T cells (T_RM_) persisting in the lung allograft diminishes^11^. T_RM_ are memory T cells that persist in either lymphoid or mucosal organs, do not re-circulate, and are poised to have a rapid effector response in the setting of secondary challenge to pathogen^12,13^. Many of these lung-infiltrating recipient T cells upregulate canonical surface markers of T_RM_ over the months following transplantation and have similar gene expression to persisting donor-derived lung T_RM_^11^. Importantly, the proportion of recipient-derived T cells in the lung allograft appears to correlate with the existence of ACR events, however, the specificity of recipient-derived T cells in the allograft has not been reported.

Using single cell RNA and T cell receptor (TCR) sequencing of recipient-derived T cells found in the bronchoalveolar lavage (BAL) of patients with ACR and after successful treatment with methylprednisolone, we set out to determine whether recipient-derived T cells recruited to the lung allograft during ACR persisted as T_RM_. Furthermore, we sought to compare gene expression of persisting recipient T cell clones before and after treatment with glucocorticoids. We found that at the time of ACR, the lung allograft contains clonally expanded population of cytotoxic and effector CD8^+^ T cells that universally persist after successful treatment with glucocorticoids. These expanded clones are composed predominantly of effector memory T cells with rapid upregulation of gene and protein expression of canonical markers of tissue residency. Finally, we show that these clones are found to aggregate around the airways consistent with lymphocytic bronchitis, the pathologic precursor of bronchiolitis obliterans, the major histologic feature of CLAD.

## Results

### Phenotype and localization of recipient-derived T cells during ACR

We first set out to determine the location of recipient-derived T cells in the lung allograft at the time of ACR and the phenotype of recipient-derived T cells found in the allograft at the time of ACR and compare this with those found before ACR and after successful treatment with high-dose glucocorticoids. To accomplish this, we identified a cohort of 15 double lung transplant recipients for whom we had cryopreserved cells obtained from the BAL at the time of ACR and at least one sample from either before ACR or after successful treatment (**Table 1**). Most study participants had formalin-fixed paraffin embedded (FFPE) transbronchial biopsy specimens stored from the same time as BAL acquisition. We performed immunofluorescence imaging of CD3 and recipient human leukocyte antigen (HLA) on FFPE transbronchial biopsies as well as multiparameter spectral flow cytometry characterization of recipient-derived T cells found in the BAL before, during, and following successful treatment for ACR. In both biopsies and BAL, recipient derived T cells were isolated by staining for recipient-specific human leukocyte antigens (HLA). From transbronchial biopsies clinically determined to have ACR, we found that perivascular lymphocytic infiltrates consisted mainly of recipient derived CD3^+^ lymphocytes, with some donor-derived CD3^+^ lymphocytes seen within the parenchyma, more removed from the vascular space (**Figure 1A**).

**Figure 1.**
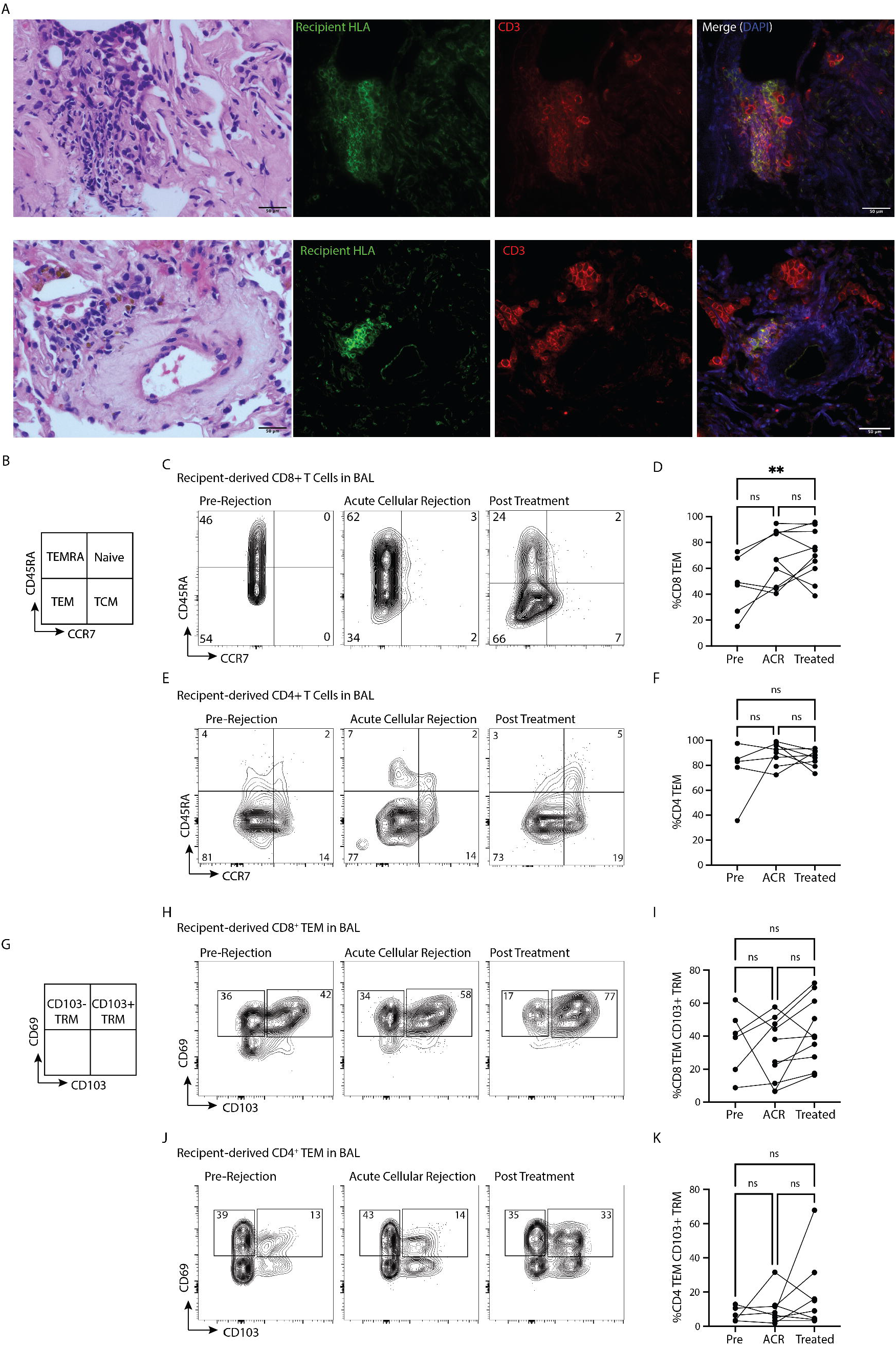
Allograft infiltration by recipient-derived effector memory T cells during ACR. (A) Immunofluorescence imaging of transbronchial biopsies from one study participant (P16) with ACR (top) and after treatment (bottom); green represents recipient-derived HLA (HLA-B7), red represents CD3. Perivascular clusters of recipient-derived T cells (yellow cells on merged image) are seen in both images, but to a lesser extent after treatment. (B) Definition of T cell phenotypes based on CD45RA and CCR7 expression. (C) Representative flow cytometry of CD8^+^ T cell phenotype from BAL pre (left), during (center), and after (right) ACR. (D) Cumulative data of CD8^+^ T cell phenotype (N=10, **p-value < 0.01). (E) Representative flow cytometry of CD4^+^ T cells from BAL pre (left), during (center), and after (right) ACR. (F) Cumulative data of CD4^+^ T cell phenotype (N = 10, ns = no statistically significant difference). (G) Defining different phenotypes of T_RM_ based on CD69 and CD103 expression. (H) Representative flow cytometry of CD8^+^ T_RM_ from the BAL pre (left), during (center), and after (right) ACR. (I) Cumulative data of CD8^+^ T_RM_ (N = 10). (J) Representative flow cytometry of CD4^+^ T_RM_ from the BAL pre (left), during (center), and after (right) ACR. (K) Cumulative data of CD4^+^ T_RM_ (N = 10).

We next performed recipient T cell phenotyping of serial BAL samples obtained from study participants who developed ACR. Based on cell surface CD45RA and CCR7 expression, T cells were divided into Naïve (CCR7^+^CD45RA^+^), effector memory T cells (T_EM_, CCR7^-^ CD45RA^-^), terminally differentiated effector T cells (T_EMRA_, CCR7^-^CD45RA^+^), and central memory T cells (T_CM_, CCR7^+^CD45RA^-^) (**Figure 1B**). We found that the proportion of CD8^+^ T_EM_ increased over the time-course of ACR (**Figure 1C**) and that the majority of CD4^+^ T cells were composed of T_EM_ regardless of clinical state (**Figure 1D**). The two most reported canonical cell surface markers for TRM in humans are CD69^14^ and integrin alpha E (CD103)^15^, with CD103 expression highlighting a subset of T cells that have a particularly rapid and robust effector response to secondary challenge with inhaled pathogens^15^. We found no change in the BAL content of either CD4^+^ or CD8^+^ T_RM_ over the course of ACR (**Figure 1E-1G**).

### Clonally expanded T cells found in the BAL at the time of ACR invariably persist as T_RM_

To determine if there is T cell clonal expansion within the lung allograft at the time of ACR, and if these expanded clones persist as T_RM_, we performed single cell RNA and T cell receptor (TCR) sequencing of FACS-sorted recipient-derived T cells found in the BAL of three study participants at the time of ACR and after successful treatment with systemic high-dose glucocorticoids (as well as one early sample predating ACR, **Figure 2A**). Any T cell clone persisting within the allograft across two timepoints was determined to be a definitive T_RM_. We identified samples from two study participants with late (> 1 year) ACR and one from intermediate (6-12 month) ACR. Late ACR samples were obtained at 17 months and 26 months after transplantation, intermediate ACR was obtained at 10 months after transplantation; follow up BAL with associated biopsy showing clearance of ACR was obtained at 2 months, 6 months, and 2 months after ACR, respectively. Expanded clones were defined as any TCR clone (either TCR A/B pair or orphaned TCR B) that consisted of >1% of the total recipient TCR repertoire at the time of BAL. We found an oligoclonal T cell expansion in all 7 samples, regardless of timeframe and presence of ACR (**Figure 2B-2D, Suppl fig 1**). In study participant P8, for which we had an earlier timepoint, we found that the majority of expanded clones at the time of ACR were not present at earlier timepoints (**Figure 2B**). Importantly, in all three study participants, each expanded clone at the time of ACR persisted as a T_RM_ after successful treatment systemic high-dose glucocorticoids (**Figure 2B-2D**).

**Figure 2.**
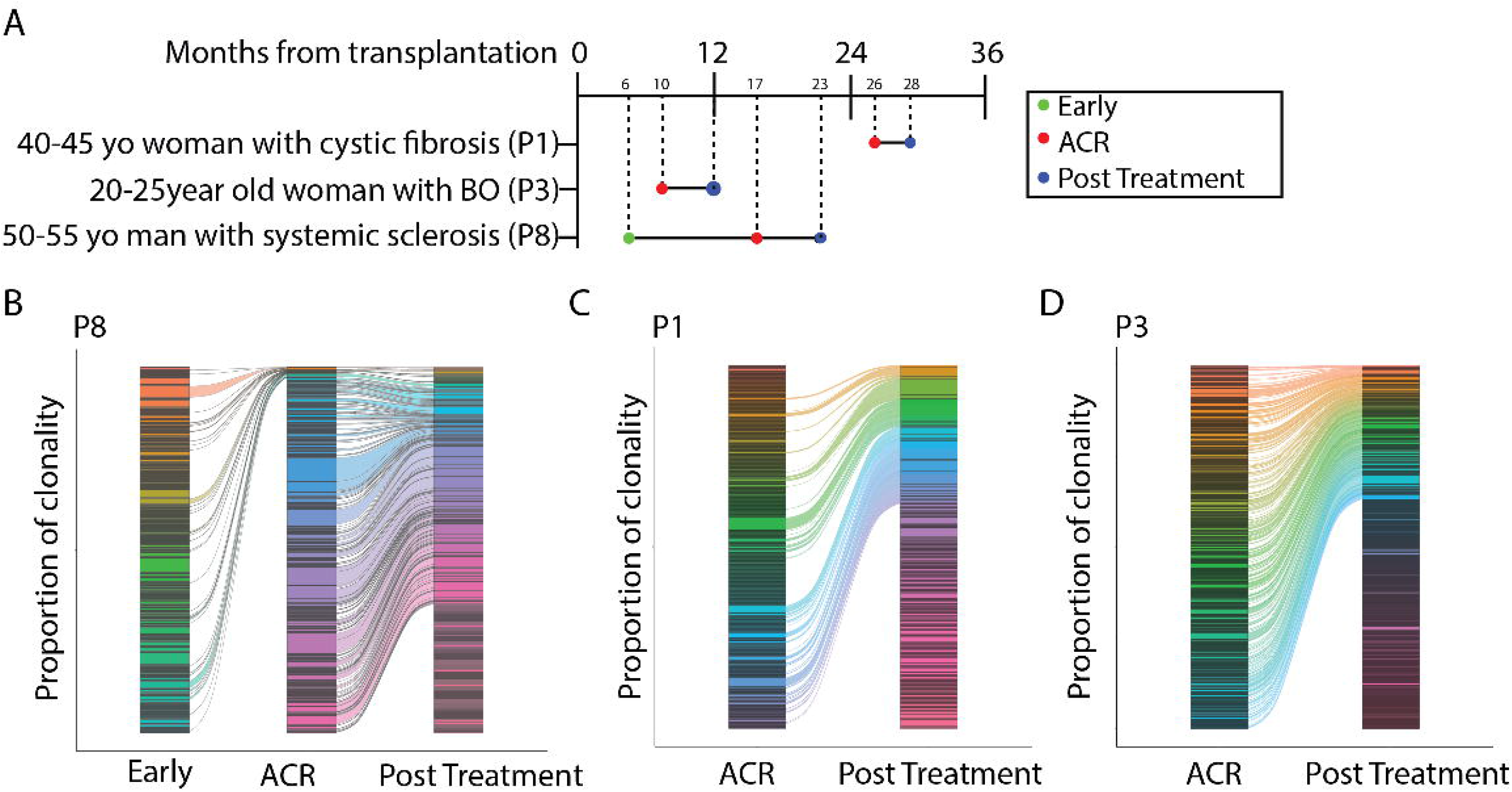
Clonally expanded recipient-derived T cells at the time of ACR persist as T_RM_. (A) Experimental design outlining longitudinal sampling for single-cell RNA and TCR sequencing of recipient-derived T cells found in the BAL of lung transplant recipients. (B) Recipient-derived T cell clonal overlap form longitudinal BAL samples obtained from three study participants, P8 (left, with three timepoints at 6 months, 17 months, and 23 months after transplantation, P1 (center, with two timepoints at 26 months and 28 months after transplantation), and P3 (right, with two timepoints at 10 and 12 months). Each color represents a unique clonotype; TCRA/TCRB pairs and orphaned TCRB are included separately in the analysis.

### Clonal expansion of cytotoxic CD8^+^ T_RM_ in the BAL during ACR

We next sought to determine the transcriptional phenotype of expanded clones at the time of late (> 2 years after transplantation) ACR using combined single cell RNA and TCR sequencing of recipient-derived T cells obtained at the time of ACR and after successful treatment. From the BAL of participant P1 (obtained at 26 months after transplantation at ACR and 28 months at time of treatment) we identified 10 distinct clusters of recipient-derived T cells based on single-cell transcriptional profiling (**Figure 3A**). Two distinct populations of clusters were identified, with the smaller population containing cells with an effector-memory gene expression profile and a larger population of clusters with gene expression more consist with naïve cells (based on *S1PR1* and *SELL* expression). The majority of T cell clonal expansion at the time of ACR was limited to the two most populous clusters within the UMAP (**Figure 3B, left**). Following successful treatment of ACR with high-dose systemic glucocorticoids, clonal expansion persisted in, but was not limited to, the original two highly expanded clusters (Fig 3B, right). When highlighting the top 4 expanded clones within the UMAP we found shared expansion of clones between both clusters 0 and 1, all of which remain within the same cluster after treatment (**Figure 3C, Suppl fig 2**). When highlighting the top 4 clones expanded at the time of treatment, we found two clones persisting within clusters 0 and 1, as well as expansion of clones shared between clusters 3 and 4 (**Figure 3D, Suppl fig 2**). Overall, most expanded clones stayed within the same gene expression cluster, regardless of presence or absence of ACR.

**Figure 3.**
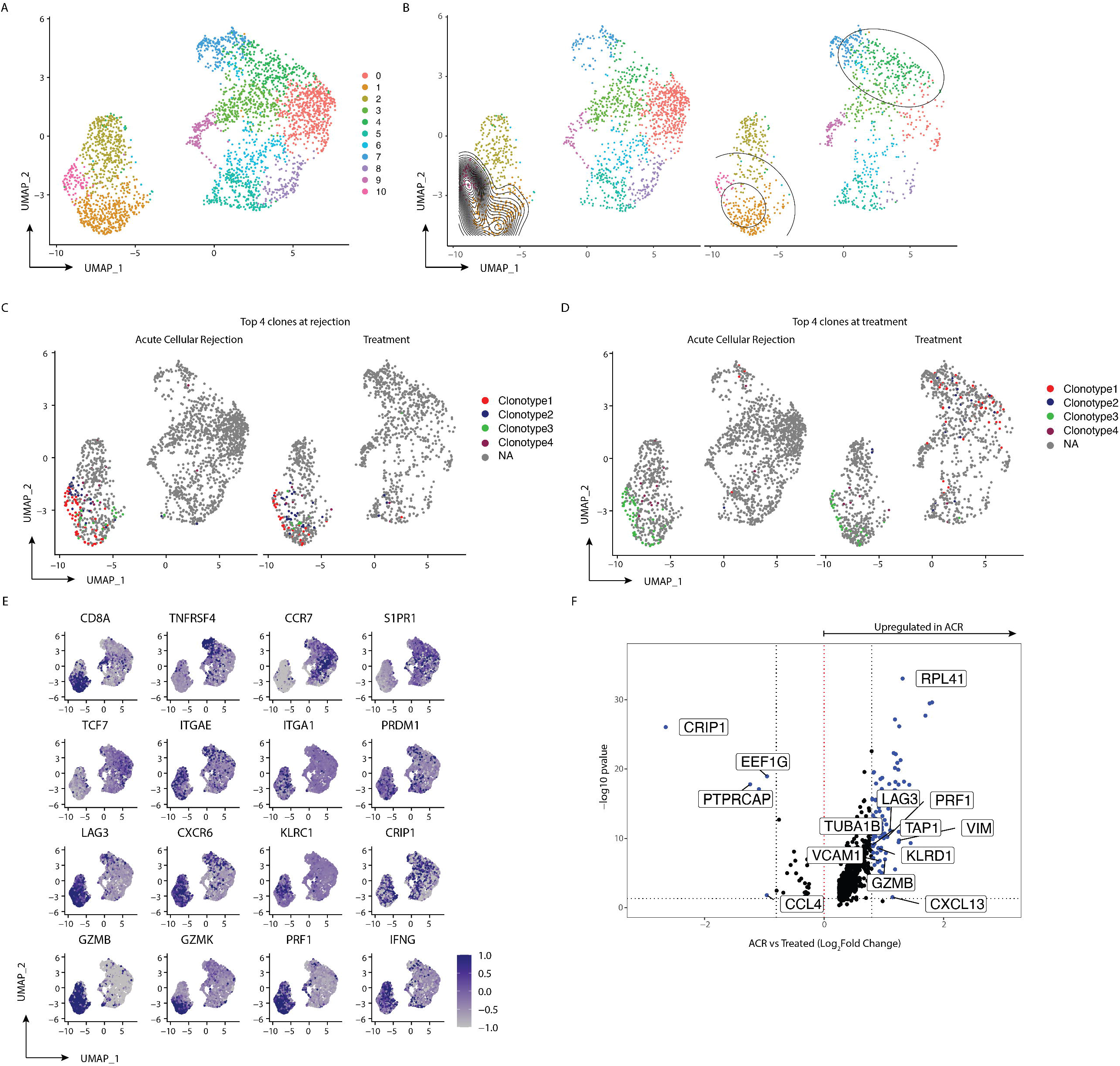
Clonal expansion of cytotoxic CD8^+^ T cells during ACR that persist as T_RM_. (A) UMAP of concatenated samples from study participant P1 at the time of ACR and after treatment showing 10 distinct clusters of recipient-derived T cells. (B) Density plot of clonal expansion overlaying UMAP, separated by cells obtained at the time of ACR (left) and after treatment (Right). (C) UMAP separated by ACR (left) and treatment (right) samples highlighting the top 4 clonotypes present at the time of ACR (paired TCRA/TCRB and orphaned TCRB are combined by shared TCRB). (D) UMAP separated by ACR (left) and treatment (right) samples highlighting the top 4 clonotypes present at the time of treatment (paired TCRA/TCRB and orphaned TCRB are combined by shared TCRB). (E) Feature plots of concatenated samples from participant P1. (F) Volcano plot showing differential gene expression between ACR and treatment using subset of top 4 clones at the time of ACR.

The two clusters containing the highest degree of T cell clonal expansion at the time of ACR had increased expression of *CD8A* (**Figure 3E**). Additionally, they had high expression of genes associated with cytotoxicity (*GZMB, GZMK, PRF1*) and effector function (*IFNG*). Interestingly, these expanded clones had high increased expression of genes related with tissue residency (*ITGAE, ITGA1, PRDM1, CXCR6, LAG3*) and downregulation of genes associated with tissue egress (*CCR7, S1PR1*), suggesting an early transcriptional signature favoring tissue retention. Interestingly, we found upregulation of *KLRC1*, the natural killer cell inhibitory receptor, which, in the context of viral infections, works to diminish viral-specific cytotoxicity^16^. Clusters 3 and 4, displayed more clonal expansion at the time of successful treatment, had genes upregulated for *TNFRSF4*, suggesting CD4+ T cells, as well as *CCR7* and *S1PR1*, consistent with a naïve phenotype (**Figure 3E**). When we performed subset analysis of the top 4 clones at the time of ACR and compared gene expression from these persisting clones between ACR and treatment, we found that these expanded clones downregulated genes related to cytotoxicity (*GZMB* and *PRF1*) (Figure 3F). The most upregulated gene after treatment was *CRIP1*. Together these findings show that during ACR, the allograft contains a clonal expansion of cytotoxic, recipient-derived CD8^+^ T cells, which all persist as T_RM_ after successful treatment of clinical ACR with high-dose systemic glucocorticoids, but with reduced expression of functional markers of cytotoxicity.

### Differential clonal expansion in the allograft versus the circulation

One prior study in kidney transplant recipients showed a shared clonal expansion in the renal allograft and circulation during acute cellular rejection^17^. To determine if this relationship was true in the lung allograft, we compared the TCR repertoire from the BAL of study participant 2 with the bulk TCR Beta chain repertoire obtained from circulating recipient-derived T cells at 3 months, 9 months, and at the time of ACR (16 months) after transplantation. From the BAL, we found a clonal expansion of cytotoxic CD8^+^ T_RM_, similar to findings in study participant 1 (**Figure 4A, 4B**). Like participant 1, all expanded clones persisted, albeit at reduced frequencies, following successful treatment of ACR with high-dose methylprednisolone (**Figure 4C**).

**Figure 4.**
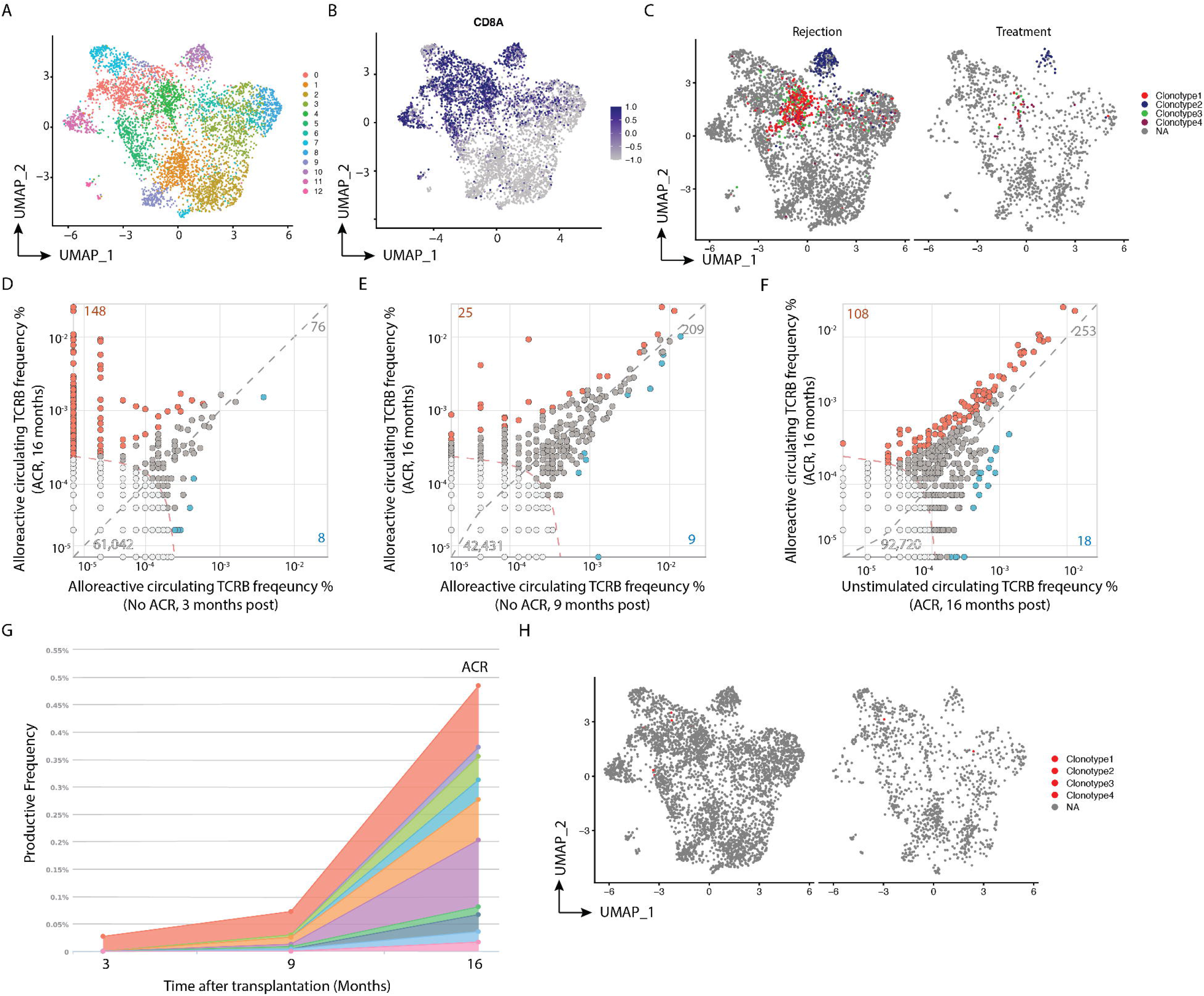
T cell clonal expansion is discordant between allograft and circulation at ACR. (A) UMAP of concatenated samples from study participant P8 at the time of ACR and after treatment showing 12 distinct clusters of recipient-derived T cells. (B) Feature plot highlighting CD8^+^ T cells. (C) UMAP separated by ACR (left) and treatment (right) samples highlighting the top 4 clonotypes present at the time of treatment (paired TCRA/TCRB and orphaned TCRB are combined by shared TCRB). (D-F) Scatterplots of Bulk TCRB sequencing from DNA extracted form FACS-sorted circulating T cells obtained from study participant P8 at 3 months, 9 months, and 16 months (at time of ACR) after lung transplantation. For each scatterplot, orange datapoints represent alloreactive clones expanded at the time of ACR, blue datapoints represent clones less prevalent at the time of ACR, and grey datapoints represent clones expanded in both groups. Any datapoint on the X or Y axis represents a unique clone to that sample. (D) Alloreactive clones at the time of ACR (y-axis) compared to alloreactive clones found at 3 months (x-axis). (E) Alloreactive clones at the time of ACR (y-axis) compared to alloreactive clones at 9 months). (F) Alloreactive clones at time of ACR (y-axis) compared to all clones found at time of ACR (x-axis). (G) Clonal overlap of expanded alloreactive clones from 3-16 months after transplantation. (H) UMAP separated by ACR (left) and treatment (right) samples of T cells in BAL highlighting the top expanded clonotypes present in the circulation at the time of ACR (paired TCRA/TCRB and orphaned TCRB are combined by shared TCRB).

Bulk TCR B sequencing was next performed from DNA isolated from FACS-sorted circulating recipient-derived T cells from this same transplant recipient at different timepoints, including at the time of ACR. This was performed both on all T cells, and on CD69+ and/or CD137/CD40L^+^ T cells after a 12-hour mixed lymphocyte reaction with irradiated donor cells. CD69^+^ and/or CD137^+^ cells were labeled allo-specific. We found a polyclonal population of expanded alloreactive T cells at the time of ACR (16 months), most of which were not present at 3 months after transplant (**Figure 4D**), but most of which were already present by 9 months of transplant (**Figure 4E**). Most of the expanded circulating T cell clones at the time of ACR were alloreactive (**Figure 4F**). From the top expanded clones in the circulation, only a fraction were identified within the BAL, and when present were not clonally expanded (**Figure 4H**).

### Expanded clones migrate to the airways

To determine the anatomic localization of expanded T cell clones we used a BaseScope^™^ *in-situ* hybridization assay (Advanced Cell Diagnostics) with an RNA probe for the CDR3β region of the top expanded clone identified in the BAL of participant 2 at the time of ACR. We performed in-situ hybridization using this probe on formalin fixed paraffin embedded transbronchial biopsies obtained from participant 2 at four different time points, two preceding ACR, one at the time of ACR, and once after successful treatment with methylprednisolone. The clone of interest was not visualized in the two samples obtained prior to developing ACR. At the time of ACR, the clone was identified within the lung parenchyma (**Figure 5A**). Following successful treatment of ACR with methylprednisolone, the same clone was identified, this time within the intra-epithelial and subepithelial sections of the airway (**Figure 5B**). This suggests that expanded T cell clones found in the lung at the time of ACR migrate to the airways and persist as T_RM_.

**Figure 5.**
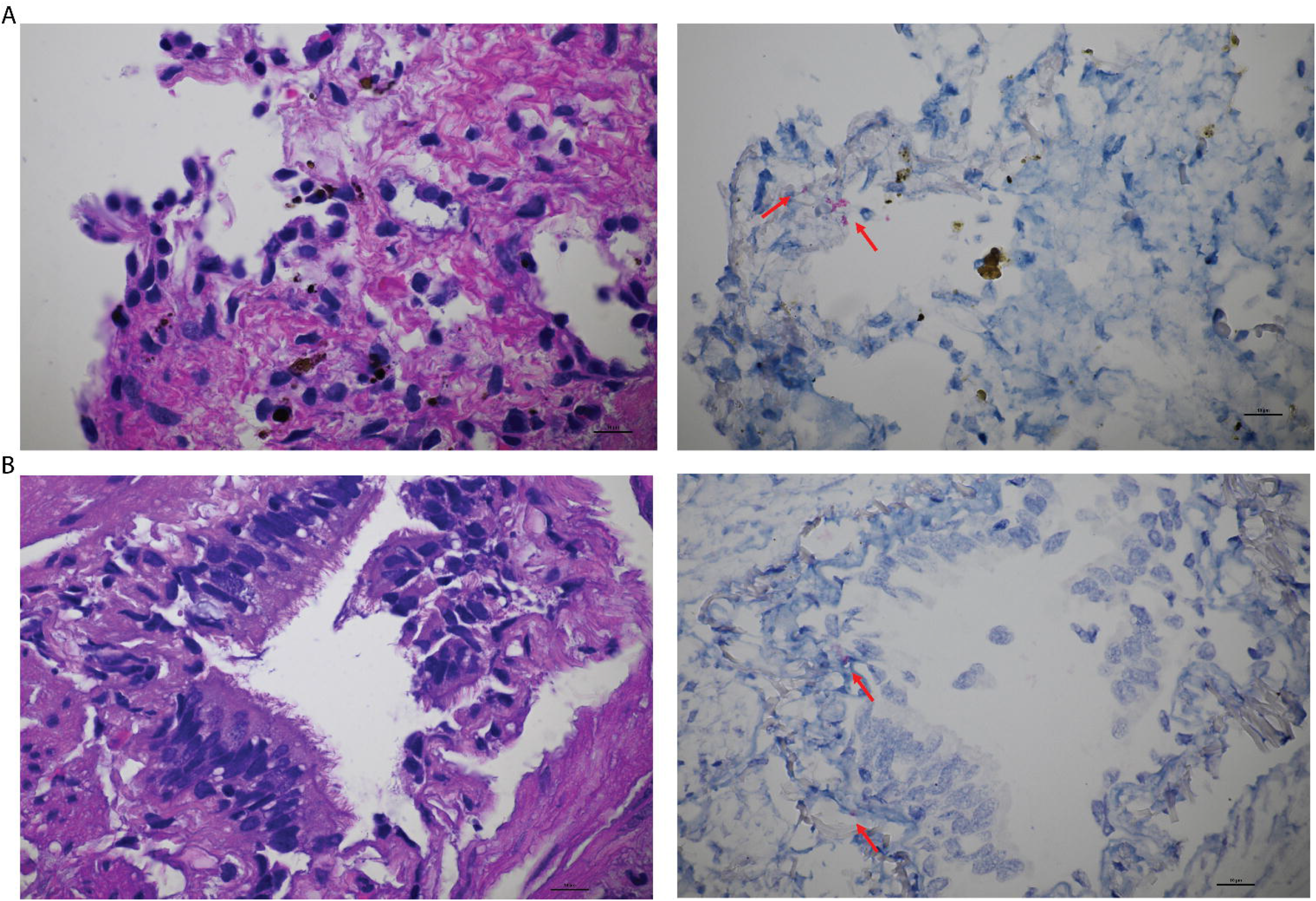
Top expanded clonotype at ACR persists in the airway as TRM. (A) Hematoxylin and Eosin (H&E) stain of transbronchial biopsy of study participant P8 at the time of ACR (left) and RNA in-situ hybridization (right, BaseScope^™^) highlighting the parenchymal presence of the top expanded clone found in the BAL at the time of ACR. (B) H&E stain of transbronchial biopsy of study participant P8 after successful treatment of ACR (left) and RNA in-situ hybridization (right, BaseScope^™^) highlighting the intraepithelial and subepithelial (arrows) presence of the top expanded clone found in the BAL at the time of ACR.

### Transcriptional re-programming of persisting clones after systemic glucocorticoid therapy for ACR

We next set out to identify the transcriptional signature of expanded clones in relation to non-expanded clones at the time of ACR and in relation to the same clones after successful treatment of ACR in concatenated samples of all three study participants. First, we identified the top 4 expanded clones from all three study participants at the time of ACR (**Figure 6A**). Comparing gene expression in the expanded clones compared to all other T cells found in the BAL, we found expanded clones had upregulation of genes associated with cytotoxicity (*GZMB, GZMH, PRF1, NKG7*), genes associated with leukocyte migration (*CCL5, XCL1, XCL2*), cellular activation (*HLA-DRB1, ZNF683*), tissue residency (*ITGAE*), and cellular exhaustion (*LAG3*). Genes downregulated in expanded clones included those related to tissue egress (*CCR7*), and one gene associated with cytotoxicity (*KLRB1*) (**Figure 6B**).

**Figure 6.**
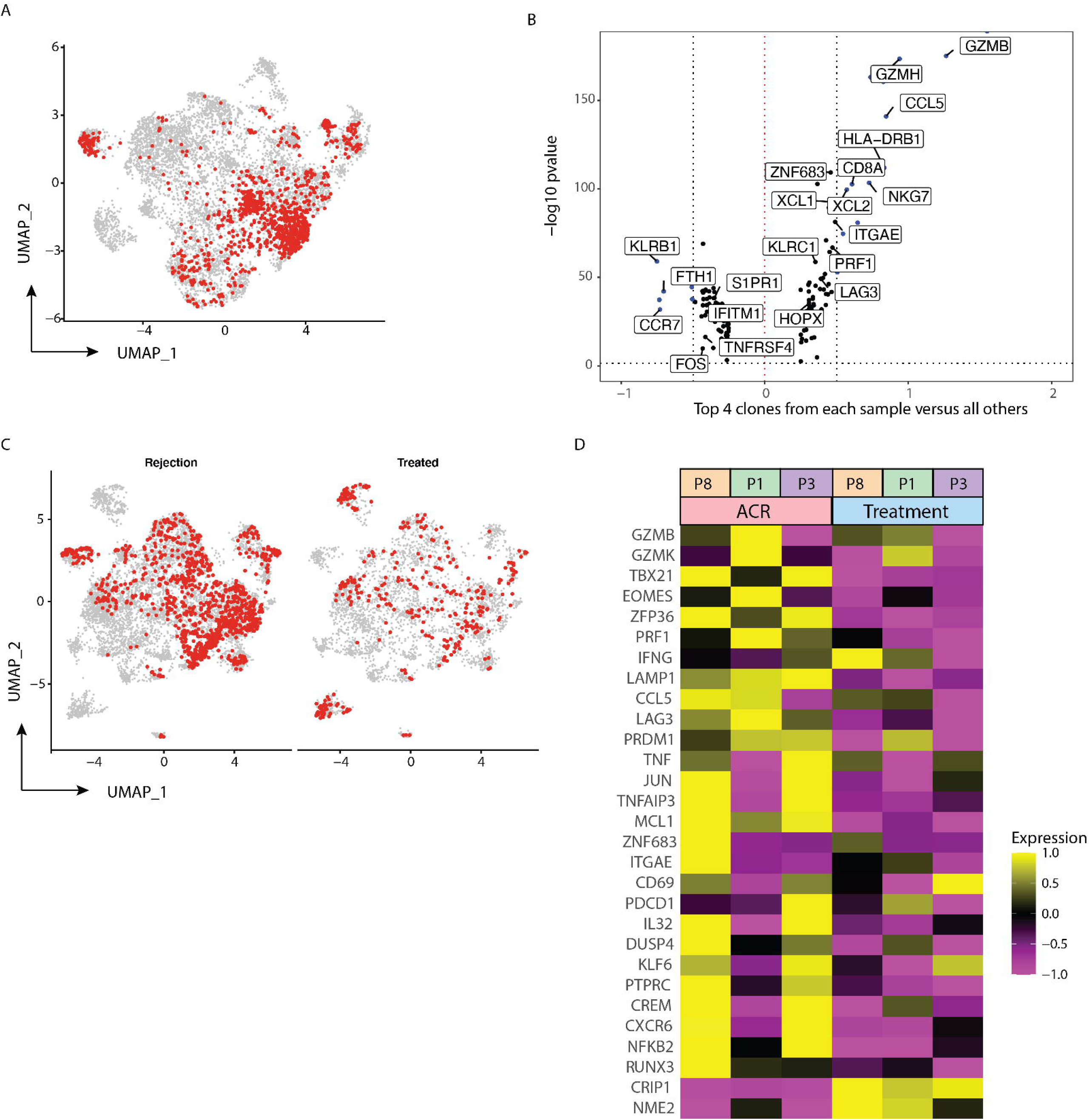
Transcriptional reprogramming of lung TRM after high-dose systemic glucocorticoids. (A) UMAP of concatenated samples from three study participants P8, P1, and P3 at the time of ACR. Red cells represent top 4 clones from each sample. (B) Volcano plot showing differential gene expression between top 4 clones from each sample at the time of ACR compared to all other clonotypes found at the time of ACR. (C) UMAP of concatenated samples from both ACR and treatment samples from three study participants P8, P1, and P3 split by presence of ACR (left) and samples obtained at time of successful treatment (right). Red cells represent top 4 clones from each sample. (D) Heatmap over averaged gene expression comparing the top 4 clones at the time of ACR for all three samples to those same clones at the time of treatment.

Next, we focused our analysis on the top 4 clones identified from each BAL sample at the time of ACR and compared gene expression from those 12 clones at the time of ACR with the same clones identified after successful treatment with methylprednisolone (**Figure 6C**). Comparing these same clones before and after treatment we found upregulation of genes at the time of ACR related to cytotoxicity (*GZMB, GZMK, PRF1, LAMP1*), effector function (*IFNG, TNF*), cellular exhaustion (*PDCD1, LAG3*), and transcription factors associated with effector function (*TBX21, EOMES*). The two most upregulated genes after successful treatment were *CRIP1* and *NME2* (**Figure 6D**). These findings support that allograft-persisting CD8^+^ T_RM_ are transcriptionally re-programmed following clinical clearance of ACR with high-dose, systemic glucocorticoids. It remains unknown if this is an effect, either direct or indirect, of glucocorticoid therapy, or whether this is a conditioned response from local drivers or persistent antigen exposure.

### Production of cytotoxic mediators correlates with transcriptional profile

Transcriptional analysis of recipient T cells found in the BAL at the time of ACR clearly identify a clonally expanded population of CD8^+^ T cells with a transcriptional profile suggesting a cytotoxic T_RM_ phenotype. To determine if the protein production of recipient derived T cells in the BAL during ACR correlates with gene expression, we performed multiparameter flow cytometry on unstimulated cells derived from 5 patients at the time of ACR and compared the results with BAL T cells obtained from the same 5 patients after successful treatment of their ACR. A total of 36,521 live, recipient-derived CD3^+^ T cells from ten samples obtained from 5 study participants (range of 1,945 – 16,391 cells per participant) were included in a concatenated t-distributed neighbor embedded (tSNE) 2-dimensional reduction of protein expression. There was a near equal proportion of CD8^+^ and CD4^+^ T cells found in the BAL, the majority of which were TEM, with one cluster of cells found predominantly during ACR (**Figure 7A**). All study participants had T cells within the ACR enriched cluster, but to varying degrees, with three participants having a much greater proportion (**Suppl fig 3**). The cluster of cells relatively unique to ACR were found to have increased cell surface expression of CD69, but not CD103 (**Figure 7B**). These cells had low expression of Ki67 suggesting they were not proliferating. This same cluster of ACR-specific cells had high quantity of mediators of cytotoxicity (Granzyme B, Granzyme K, Perforin) as well as increased surface expression of KLRC1, a protein believed to be instrumental in negative feedback of NK-cells (**Figure 7C**). Immunofluorescence imaging of transbronchial biopsies obtained at the time of ACR show that pathognomonic lesions for ACR (perivascular infiltrate of lymphocytes) were predominantly composed of recipient-derived T cells with high expression for Granzyme B (**Figure 8A)** with airway-centered T cells with similarly high content of granzyme-B (**Suppl fig 4A**).

**Figure 7.**
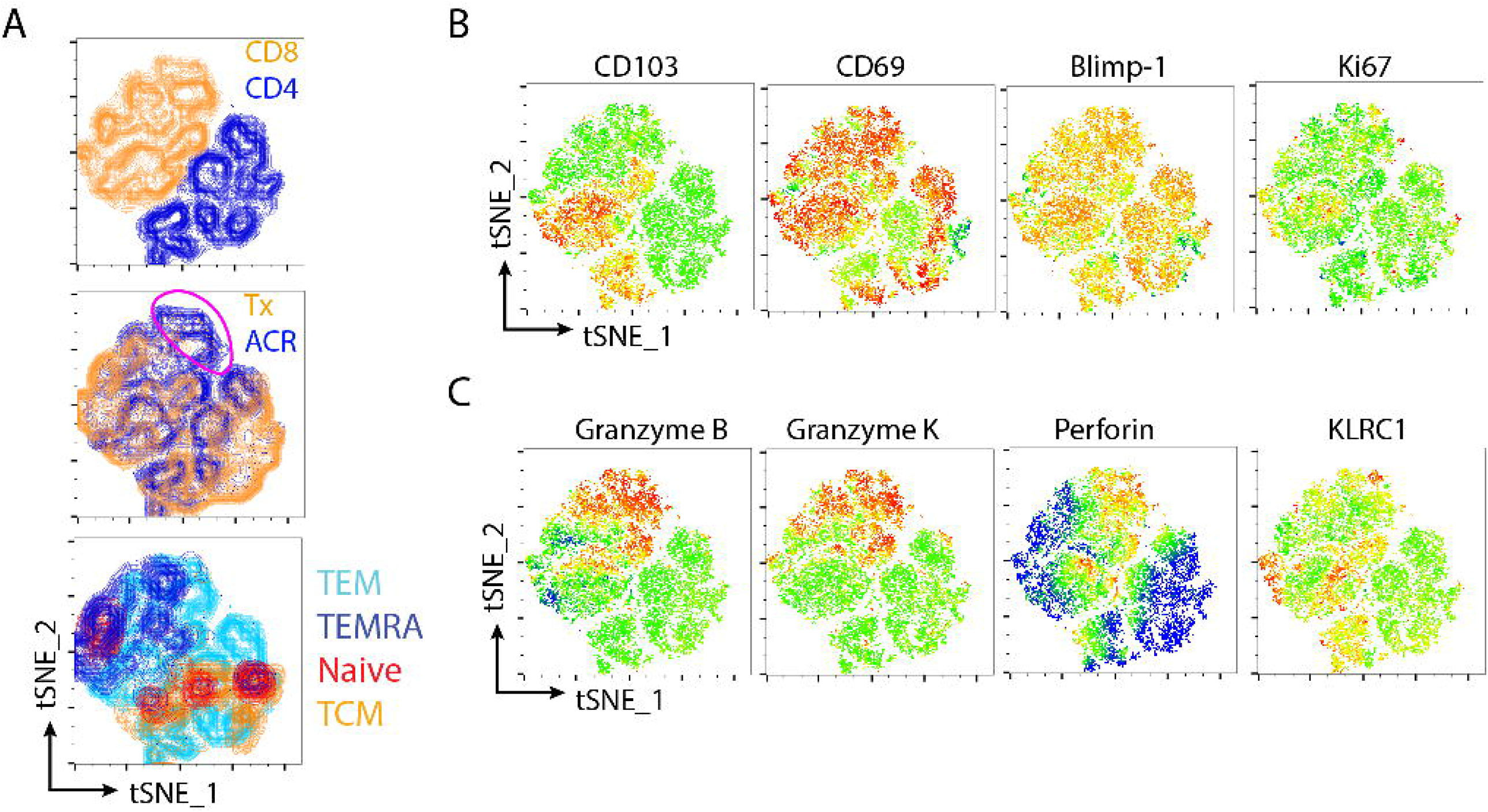
Recipient T cells at ACR have high content of cytotoxic mediators. (A-C) t-distributed stochastic neighbor embedding (tSNE) of 10 concatenated flow cytometry samples obtained from 5 study participants (each sample has one at the time of ACR and one after successful treatment), including 36,521 total cells. (A) tSNE showing T cell phenotype from 10 samples (encircled cluster in middle plot highlights ACR-enriched cluster of CD8+ T_EM_). (B) tSNE highlighting proteins related to tissue residency (CD103, CD69, Blimp-1), and proliferation (Ki67). (C) tSNE highlighting cytotoxic mediators (Granzyme B, Granzyme K, Perforin), and presumed self-regulatory protein KLRC1.

**Figure 8.**
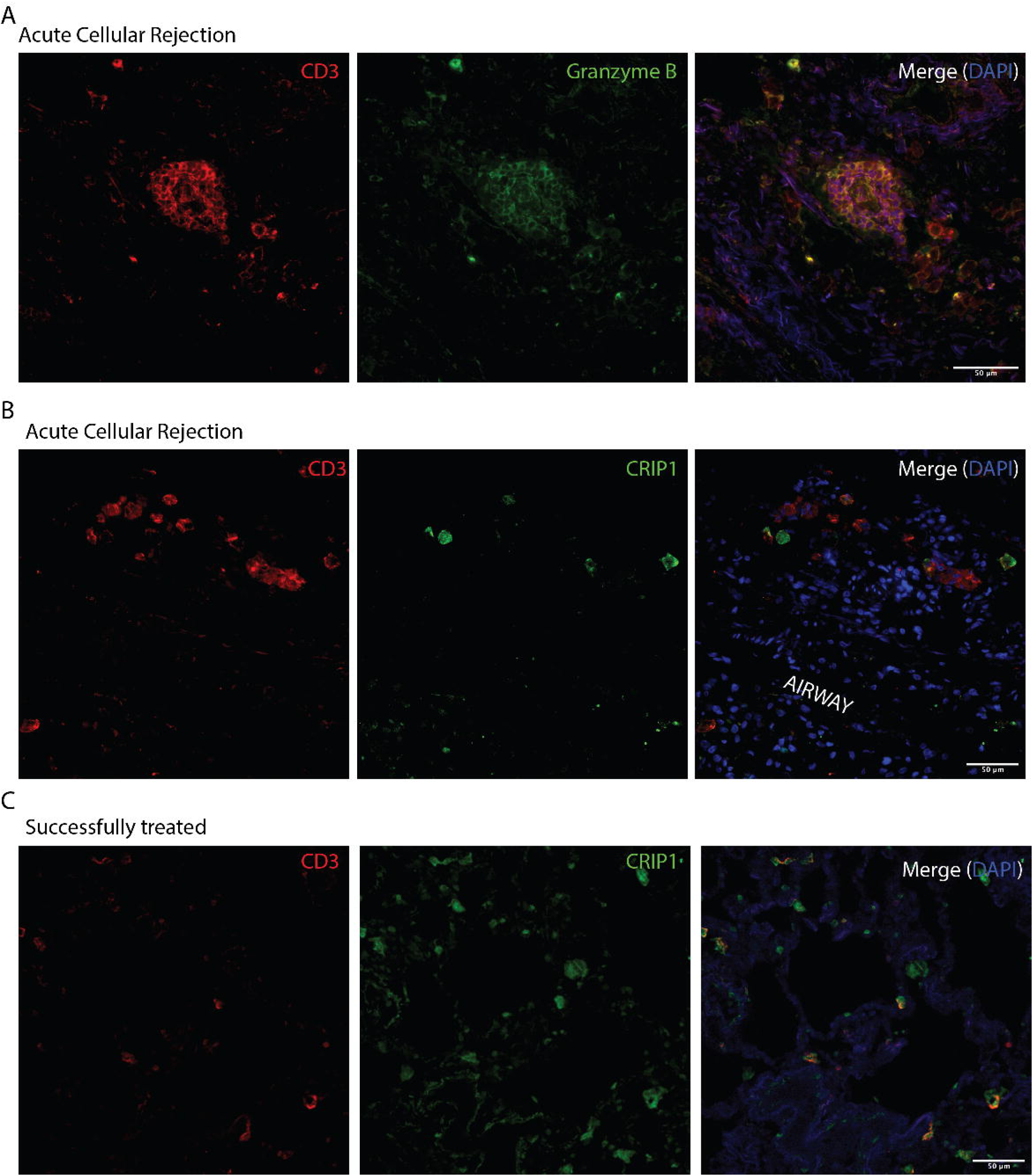
Anatomic localization of cytotoxic T cells and CRIP1 co-localization after treatment. (A) Immunofluorescence imaging of CD3 (red) and granzyme B (green) highlighting a perivascular infiltrate of double positive cells at the time of ACR. (B) CRIP1 protein content (green) does not co-localize with CD3 (red) at the time of ACR. (C) CRIP1 co-localizes with CD3 (red) after successful treatment with high-dose glucocorticoids.

Cysteine Rich Intestinal Protein 1 (CRIP1) is an incompletely characterized double zinc-finger LIM protein that is abundantly expressed in the intestines and in peripheral blood mononuclear cells^18^. We found that *CRIP1* was the most consistently upregulated gene in persisting recipient derived T_RM_ within the BAL. When performing immunofluorescence imaging of transbronchial biopsy specimens we found substantial but non-cellular specific content of the CRIP1 protein (Figure 8B). However, following successful treatment of ACR with methylprednisolone, we saw increased colocalization of the CRIP1 protein with lung T cells (**Figure 8C, Suppl fig 4B**,**4C**). Together these findings confirm that, during ACR, the lung contains a clonally expanded population of cytotoxic, recipient derived CD8^+^ T cells that universally persist in the lung as TRM after successful treatment with high dose glucocorticoids. Following treatment, the lung T cell population downregulates gene and protein expression of cytotoxic mediators and upregulates CRIP1.

## Discussion

Acute cellular rejection represents a major burden to patient morbidity following lung transplantation and is associated with an increased risk of early CLAD, the major limiting factor to long-term survival after transplantation. Herein, we show that ACR of lung allografts is notable for a perivascular infiltrate of recipient-derived CD3^+^ lymphocytes, most of which have an effector memory (T_EM_) phenotype followed by a terminally differentiated effector (T_EMRA_) population. From single cell RNA/TCR sequencing of recipient-derived T cells obtained in the BAL of three study participants with active ACR, we show that ACR is characterized by an oligoclonal CD8^+^ T cell expansion within the allograft, with expanded clones having a cytotoxic gene expression and protein production, dominated by granzyme B, granzyme K, and perforin. We found that, in all three study participants, all expanded clone (>1% of the total clonal population) persisted as T_RM_ weeks to months after successful treatment with systemic, high-dose glucocorticoids, however, with a reprogrammed transcriptional profile. The most highly expanded clone identified, was noted on biopsy to migrate to the airways after treatment. Finally, we show that clonal expansion within the allograft was discordant with oligoclonal expansion in the periphery at the time of ACR. Together these findings suggest that cytotoxic T cells recruited to the allograft during ACR develop into lung T_RM_ that persist despite systemic high-dose glucocorticoids.

We previously reported that over the months following lung transplantation, graft-infiltrating, recipient-derived T cells develop a phenotype of tissue residency, upregulating proteins that promote tissue retention^11^. These include CD69 which promotes downregulation of the protein S1PR1 thereby diminishing lymphatic egress^19^, CD103 (integrin alpha E) which binds to E cadherin promoting retention near epithelial cells^20,21^, and CD49 (integrin alpha 1) which promotes retention by binding to collagen^22,23^. Importantly, this accumulation of recipient derived T cells in the allograft occurred faster in the setting of ACR^11^. In a murine model of delayed rejection after orthotopic renal transplantation, polyclonal, antigen-specific CD8^+^ T_RM_ populated the graft, and contributed to the development of chronic rejection^24^. Furthermore, renal allografts after human transplantation contained CD8^+^ T_RM_ that could produce large amounts of granzyme B, perforin, IFNγ, and TNFα after stimulation with the phorbol ester, PMA/Ionomycin^25^. Our finding that clonally expanded CD8^+^ T cells found during ACR persist as T_RM_ and migrate to the airways suggests a plausible biologic mechanism whereby ACR contributes to the bronchiolitis obliterans syndrome (BOS) phenotype of CLAD. Indeed, we recently demonstrated a strong Type-1 immunity gene signature using bulk RNAseq obtained from airway brushings of the distal small airways in patients with CLAD, consistent with these findings^26^. Although the T_RM_ appear to undergo transcriptional reprogramming following systemic glucocorticoid therapy, further study is required to see whether, like other TRM, they are capable of rapid re-activation.

Bulk RNA sequencing of the cellular component of BAL from lung transplant recipients with and without ACR previously identified genes related to cellular cytotoxicity to be upregulated during ACR, most notably, *GZMK* and *GZMA*, as well as effector gene *IFNG*^3^. However, despite this cytotoxic profile, we cannot say definitively that the expanded clonal population found in the allograft at the time of ACR is composed of purely alloreactive clones. The oligoclonal nature of the expansion, as well as the discordant clonal expansion from circulating T cells would suggest that this does not represent bystander activation. This discordant clonal expansion, however, does raise the possibility of local expansion of previously established TRM as a possible source, which has been shown to occur in a murine model of recurrent skin infections^27^. However, this may also represent bystander activation. Renal transplant studies have shown CMV-specific T cells to have cross-reactivity with the allograft^28^, or other inhaled viral pathogens or microbiome. Due to the paucity of cells identified, we were unable to do proliferation studies on these samples to definitively determine allo-reactive potential.

The clinical management of ACR after lung transplantation varies greatly by institution and individual provider^9^. Despite this heterogeneity in clinical approach, systemic glucocorticoids remain the standard first-line therapy for symptomatic ACR^29^. The use of glucocorticoids to treat ACR is largely based on extrapolation from kidney transplant practice and serial pathologic assessments of lung allografts showing diminished perivascular infiltration after treatment^30^. The impact of systemic glucocorticoids on diminishing ACR has been postulated as an effect on circulating T cells, both diminishing the number of alloreactive cells via apoptosis^31^, and reducing the activation and cytokine production of alloreactive cells via inhibition of interleukin 2 (IL2) signaling and production^32^. T_RM_ represent a unique subset of memory T cells that are relatively removed from the circulation, even in the highly vascular human lung^33^. This sparing from the circulation is believed to be the reason why T_RM_ are relatively protected from the effects of some systemic lympho-depletive therapies^34^. The ability of systemic glucocorticoids on mucosal T_RM_ has yet to be reported. Here, we show a transcriptional reprogramming of lung T_RM_ after the administration of high dose glucocorticoids, suggesting an impact on local resident immunity. It remains unclear whether this is a direct effect of glucocorticoids on the TRM, or an indirect effect via circulating impaired helper T cells^35^, regulatory T cells^36^, or through impacts on the local environment. Further study is required to elucidate how systemic glucocorticoids impact mucosal T_RM_ function. The role of CRIP1 expression after successful treatment is similarly unknown. CRIP1 is a zinc-binding protein with high expression in immune cells and epithelium that may play a role in DNA damage repair^18,37,38^. Its role in allograft recovery after ACR requires further study.

In conclusion, we show that during ACR, the human lung allograft contains a clonally expanded population of recipient-derived CD8^+^ T cells that persist as transcriptionally-reprogramed T_RM_ following systemic therapy with high-dose glucocorticoids.

## Methods and Materials

### Study participants

We identified a convenience sample of consecutive adults who underwent a first lung transplantation at the University of Pittsburgh between June of 2015 and July of 2018, who had consented to our IRB-approved biorepository, had cryopreserved cells obtained from BAL at the time of ACR and after successful treatment for rejection, and who had donor and recipient HLA discrepancies amenable to differentiation with commercially available flow cytometry antibodies^11^ (**Suppl Table 1**). The biorepository was approved by our institutional review board. ACR was defined clinically, as a perivascular infiltration of lymphocytes found on transbronchial biopsy.

### Sample collection, processing, and flow cytometry

BAL samples were centrifuged, and the pellet was re-expanded in fetal bovine serum (FBS) with 10% DMSO and frozen in liquid nitrogen for storage. Cells were thawed with warmed media (RPMI + 10% FBS) and strained sequentially through a 100 mm and 70 mm filter. Samples were then washed with FACS buffer (PBS + 1% FBS) and re-expanded with FACS buffer + 5 mm Fc receptor blocking solution (Human TruStain FcX^™^, Biolegend) for 10 minutes at room temperature. Afterwards, cell surface antibodies were applied at room temperature for 30 – 60 minutes and fixed on ice for 60 minutes (eBioscience Cat # 88-8824-00). For panels with intracellular staining, cells were washed with and stained in the presence of a permeabilization buffer. Flow cytometry was performed using a spectral flow cytometer (CyTek Aurora^™^) and data was analyzed using FlowJo. Methods for distinguishing donor versus recipient origin of immune cells from the BAL has been previously reported^1^. All antibodies used for flow cytometry and imaging can be found on **Suppl Table 2**.

### Immunohistochemistry and imaging analysis

Five μm sections of paraffin-embedded transbronchial biopsies were obtained from paraffin-embedded blocks maintained by the Pathology Department at the University of Pittsburgh. Slide de-paraffinization was performed with X% Xylene followed by serial dilutions of ethanol. Antigen retrieval was performed at 95ºC for 20 minutes in the presence of DAKO Target Retrieval Solution, pH 9 (Agilent) using a Decloaking Chamber^™^ NxGen (Biocare Medical). Primary antibody was stained overnight on an orbital shaker at 4°C. Secondary antibody was then applied for one hour on an orbital shaker at room temperature (**Suppl table 1** includes list of all antibodies used). Slides were washed and then stained with 1x DAPI for 5 minutes. Coverslips were then mounted with ProLong^™^ Gold antifade reagent (Invitrogen) and sealed with clear nail polish. Images were then captured within 72 hours with an epi-fluorescence microscope (Nikon Eclipse Ni) and a digital camera (Hamamatsu Digital Camera C11440). ImageJ was used to qualitatively analyze images and generate TIFF files from ND2.

### Mixed lymphocyte reaction

Peripheral blood was collected in heparinized BD Vacutainer tubes and PBMC were isolated using Ficoll gradient following standard protocols. RPMI culture media supplemented with 35% fetal bovine serum and 10% DMSO were used to freeze the purified PBMC in the vapor phase of liquid nitrogen at a density of 5-10 × 10^6^ cells per ml of media. Cryopreserved donor peripheral blood mononuclear cells (PBMC) were thawed, and half were labeled with cell trace dye (CFSE) and γ-irradiated (3000 rads). The remaining donor PBMC were lysed using sonication. Single-cell suspension of BAL obtained at different timepoints (Figure 2A) were stained with cell trace dye (Cell Trace Violet). Irradiated donor PBMC were combined with single cell suspension of labeled BAL cells at a ratio of 1:1 with the simultaneous addition of lysed donor PBMC in mixed lymphocyte reaction media (AIMV with 5% human serum and PSG) for 6 hours at 37 degrees in 5% CO2 incubator^39^.

### Single cell RNA-seq/TCR sequencing

Live, recipient-derived CD3^+^ T cells were isolated from the BAL using a BD FACSAria^™^ (BD Biosciences) sorting on live, CD3^+^, CFSE (irradiated donor PBMC) negative, singlets that were positive for recipient-derived HLA. Sorted cells were loaded onto a Chromium Next GEM Single Cell 5’ v1 Chip (10x Genomics) according to the manufacturer’s guidelines at a capture rate of 5,000 cells per sample. Libraries were sequenced using the Illumina HiSeq 2500 platform. Alignment, filtering, barcode counting, and UMI counting were performed with CellRanger v5 and CellRanger VDJ. Quality metrices for each sample can be found on **Suppl Table 3**.

### Single cell RNA-seq/TCR data processing

Single cell RNA sequencing analysis was performed using Seurat 3.0 with R (version 3.6). T cell receptor repertoire data was embedded in the Seurat object metadata using scRepertoire^40^. Normalization and variance stabilization of count data was performed using scTransform^41^. Seurat objects were then integrated using 3000 identified anchors based on the previously transformed normalization values. A small number of contaminating cells of myeloid descent were removed form analyses based on CD68 expression, followed by spatial visualization of distinct clusters using Uniform Manifold Approximation and Projection (UMAP) for dimension reduction^42^. For differential expression analysis between acute cellular rejection and treatment, data was subset to include only those top 4 clones present at the time of rejection. Differential gene expression was performed form this subset using non-parametric Wilcoxon rank sum test. The results were adjusted for multiple comparisons using Bonferroni correction. T cell receptor repertoire diversity was estimated across BAL samples (**Suppl methods**). All code used for single cell analyses can be found in the following GitHub repository, including list of all R packages used for analyses: https://markesnyder.github.io/LTX_scACR/

### Bulk TCR Sequencing and analysis

Peripheral blood was collected longitudinally from lung transplant recipients as part of our ongoing transplant biorepository. Lymphocytes were isolated using density centrifugation with lymphocyte separation media (Corning® LSM). Lymphocytes were then slowly cryopreserved in fetal bovine serum with 10% dimethyl sulfoxide and stored in the vapor phase of liquid nitrogen. Samples identified for study use were slowly thawed. Genomic DNA was isolated from FACS-sorted recipient-derived circulating T cells (either allo-reactive or unstimulated) using DNeasy Blood & Tissue Kit (Qiagen). Unstimulated T cells were derived from FACS-sorting live, CD3^+^, recipient HLA^+^ lymphocytes; allo-reactive T cells were isolated by sorting CD8^+^ and CD4^+^ recipient HLA^+^ lymphocytes that were positive for activation induced markers (AIM) after a 12-hour stimulation with irradiated donor PBMC^43^. Positivity for AIM markers after mixed lymphocyte reaction were defined CD69^+^ and/or CD137^+^ for CD8^+^ T cells and CD69^+^ and/or OX40^+^ for CD4^+^ T cells (**Suppl fig 5A**). Gating strategy for sorting was established based on AIM marker expression of resting T cells in the absence of ACR (**Suppl fig 5B**). At the time of ACR, there were a substantial number of CD69^+^ recipient CD3^+^ T cells at rest (**Suppl fig 5C**). DNA was quantified with NanoDrop One (ThermoScientific). Next generation TCR-Beta sequencing of CDR3 variable region was performed using the ImmunoSeq hsTCRBkit (Adaptive Biotechnologies) and sequenced with a MiSeq 150x system (Illumina). Data was analyzed using both the ImmunoSeq Analyzer software v3.0 (Adaptive Biotechnologies)^44,45^ and Immunarch^46^.

### RNA In-situ hybridization assay (BaseScope^™^)

Transbronchial biopsy slides underwent deparaffinization, rehydration, and antigen retrieval as previously described in the *Immunohistochemistry* methods section. Slides were pre-treated with protease. Our probe, designed to bind to the hypervariable CDR3-β segment of our most abundant clone in participant 2 at the time of ACR, was hybridized to our target mRNA and amplified per the manufacturer’s instructions. Counterstaining was performed with 50% Hematoxylin solution, washed with tap water, followed by emersion in 0.02% Ammonia water and again washed with tap water prior to mounting slide. Images were captured at 60x magnification using a Nikon Eclipse Ni and a digital camera (Hamamatsu Digital Camera C11440).

### Statistical analysis

Statistical analyses were performed using R (R Foundation for statistical computing, Vienna, Austria), Python (Python Software Foundation, Fredericksburg, Virginia), and GraphPad (Prism). For all analyses, a two-tailed p-value of < 0.05 was the threshold used to determine statistical significance. IHC and IF were analyzed with ImageJ (Madison, Wisconsin). Paired t-test was used to test for difference in flow cytometry T cell phenotype before, during, and after ACR. Manuscript figures were compiled using Adobe Illustrator CC 2017 (Ventura, CA).

## Data Availability

Raw data were generated at the University of Pittsburgh and are included in the article and supplementary materials. The complete set of raw data supporting the findings of this study are available from the corresponding author MES on request.

## Abbreviations

(ACR): Acute Cellular Rejection
(CLAD): Chronic lung allograft dysfunction
(T_RM_): Tissue resident memory T cell
(MLR): Mixed Lymphocyte Reaction
(BAL): Bronchoalveolar lavage

## Acknowledgements / Funding

The authors would like to thank K. Abou-Daya at the University of Pittsburgh for guidance with single cell analysis, D. Metes at the University of Pittsburgh for guidance with mixed lymphocyte reactions, and F. Lakkis at the University of Pittsburgh for guidance and feedback. As always, a special thanks to the families of organ donors who made transplantation possible. This work was supported by a NIH K23 HL151750-01 awarded to M.E.S, and an NIH 1R01HL133184 awarded to JM.

## Disclosures

The authors declare no conflicts of interest related to this submission

## Author Contributions

MES, AB, and AC performed experiments. RL and TT oversaw and performed cell capture and library preparation for single cell RNA and TCR sequencing. KC and LI oversaw and performed read QC and alignment. EL, JP, BJ, SK, and PS assisted in sample acquisition. YZ oversaw sample storage and retrieval. HET assisted with pathology review. MES and KM analyzed scRNA/TCR and Bulk TCR data with the guidance of PS. JA assisted with RNA *in-situ* hybridization and with imaging analysis. FL, PS, and JM supervised the work and guided design. All authors provided a critical review of the manuscript and assisted with revisions.

## Notes

### Competing Interest Statement

The authors have declared no competing interest.

### Author Declarations

This study was approved by the Institutional Review Board (study number: STUDY20060250) at the University of Pittsburgh.

