## Supplementary material for "Human lung tissue resident memory T cells are re-programmed but not eradicated with systemic glucocorticoids after acute cellular rejection": Table 1

| Table 1. Study participant demographics | | | | | | |
| --- | --- | --- | --- | --- | --- | --- |
| ID | Age-range | Gender | Diagnosis | Induction | CMV (D/R) | Experiments^‡^ |
| P1 | 40-45 | F | Cystic Fibrosis | Alemtuzumab | -/- | SC, FC |
| P2 | 50-55 | F | Systemic Sclerosis | Basiliximab | +/+ | FC |
| P3 | 20-25 | F | Primary OB* | Basiliximab | +/- | SC |
| P4 | 60-65 | M | Primary OB | Basiliximab | +/- | FC |
| P5 | 60-65 | F | IPF** | Alemtuzumab | +/- | FC |
| P6 | 60-65 | M | IPF** | Alemtuzumab | +/+ | FC |
| P7 | 65-70 | F | Emphysema | Basiliximab | +/+ | FC |
| P8 | 50-55 | M | Systemic Sclerosis | Basiliximab | +/- | SC, IF, Bulk, FC |
| P9 | 65-70 | F | Emphysema | Alemtuzumab | +/+ | IF |
| P10 | 65-70 | M | IPF** | Alemtuzumab | +/+ | FC |
| P11 | 65-70 | M | Emphysema | Alemtuzumab | -/- | FC |
| P12 | 30-35 | M | Cystic Fibrosis | Basiliximab | +/- | IF |
| P13 | 50-55 | M | Silicosis | Alemtuzumab | -/+ | FC |
| P14 | 30-35 | M | GVHD^t^ | Basiliximab | -/+ | FC |
| P15 | 55-60 | M | Sarcoidosis | Alemtuzumab | -/- | FC |
| P16 | 55-60 | F | Emphysema | Basiliximab | +/- | IF |
| P17 | 30-35 | F | Cystic Fibrosis | Alemtuzumab | +/+ | IF |
| ‡SC = single cell RNA/TCR sequencing; FC = flow cytometry, IF = Immunofluorescence imaging, Bulk = Bulk TCR sequencing from PBMC  *OB = Obstructive Bronchiolitis  **IPF = Idiopathic Pulmonary Fibrosis  ^t^GVHD = Graft Versus Host Disease | | | | | | |
