## Supplementary material for "Human lung tissue resident memory T cells are re-programmed but not eradicated with systemic glucocorticoids after acute cellular rejection": Suppl methods

**Supplementary methods**

**T cell repertoire diversity**

T cell repertoire diversity within the BAL at the time of ACR and after treatment was estimated via multiple different indices. Shannon index assumes that all clones are represented in the sample and that these clones have bene randomly selected, and is =

$$-\sum_{i=1}^{c} {\frac{n}{N}}_{i}\ln{\frac{n}{N}}_{i}$$

Where c = the number of distinct clones, $\frac{n}{N}$ is the proportion of an individual clone (n) over clonal abundance (N).

Inverse Simpson index places more weight on dominant clones and =

$$1 /(1- \sum_{i=1}^{c} \left( \frac{n}{N} \right)^{2})$$

Chao1-index focuses on clonal abundance, including only those clones with a single or two copies and is =

c + α_1_ (α_1_ – 1) / (2α_2_ + 1)

Where α1 is the number of single clones and α2 is the number of clones with two copies.

ACE (Abundance-based coverage estimator) is another index of clonal richness, like Chao1, but focusing on all clones with 10 or less copies each (instead of just 1 and 2 copies) and is =

$$\sum_{i=1}^{c} p_{i}I(N_{i}>0)$$

Where p_i_ = $\frac{N_{i}}{N}$, and I(A) is the indicator function(1).
