## Supplementary material for "Human lung tissue resident memory T cells are re-programmed but not eradicated with systemic glucocorticoids after acute cellular rejection": Suppl Table 1

| Supplementary Table 1. Study participant HLA | | |
| --- | --- | --- |
| **ID** | **Recipient HLA** | **Donor HLA** |
| P1 | A3, A30, B35, B62 | A2, A11, B7, B60 |
| P2 | A1, A23, B27, B35 | A1, A24, B8, B13 |
| P3 | A31, A68, B40, B44 | A2, B42, B62 |
| P4 | A24, A26, B13, B27 | A1, A11, B37, B51 |
| P5 | A2, A29, B44, B51 | A30, A33, B13, B35 |
| P6 | A2, A23, B7, B27 | A2, A32, B35, B44 |
| P7 | A3, A33, B15, B55, B65 | A2, A24, B44, B56 |
| P8 | A2, A3, B7, B57 | A1, A74, B57, B72 |
| P9 | A3, A23, B7, B44 | A1, A24, B7, B8 |
| P10 | A2, A24, B8, B51 | A2, A3, B27, B44 |
| P11 | A28, A68, B39, B44 | A11, A30, B7 |
| P12 | A3, A24, B7 | A2, A24, B7, B44 |
| P13 | A2, B44, B51 | A3, A31, B7, B62 |
| P14 | A1, A2, B49, B56 | A2, A68, B7, B51 |
| P15 | A3, A26, B18, B38 | A1, A2, B18, B35 |
| P16 | A3, A23, B7, B18 | A24, B35, B39 |
| P17 | A24, A74, B51, B72 | A1, B8 |
