## Supplementary material for "Human lung tissue resident memory T cells are re-programmed but not eradicated with systemic glucocorticoids after acute cellular rejection": Suppl Table 2

| Supplementary Table 2: antibodies used for Experiments | | | | |
| --- | --- | --- | --- | --- |
| **Multiparameter flow cytometry and FACS Sorting** | | | | |
| **Antibody** | **Clone** | **Fluorochrome** | **Supplier** | **Catalog #** |
| Viability | NA | Zombie NIR | Biolegend | 423106 |
| Viability | NA | Zombie Aqua | Biolegend | 423102 |
| CD45 | HI30 | PE/Cy5 | Biolegend | 304010 |
| CD3 | SK7 | BUV395 | BD Biosciences | 564001 |
| CD4 | SK3 | BV480 | BD Biosciences | 566104 |
| CD8 | SK1 | BUV737 | BD Biosciences | BDB612754 |
| CCR7 | G043H7 | BV785 | Biolegend | 353230 |
| CD45RA | HI100 | PED594 | Biolegend | 304145 |
| CD69 | FN50 | BV650 | Biolegend | 310934 |
| CD103 | Ber-ACT8 | APC/Cy7 | Biolegend | 350228 |
| HLA-A2 | BB7.2 | PE | Biolegend | 343306 |
| HLA-A2 | BB7.2 | FITC | Biolegend | 343304 |
| HLA-B7 | BB7.1 | PE | Biolegend | 372404 |
| HLA-ABC | W6/32 | FITC | Biolegend | 311404 |
| HLA-B8 | REA145 | PE | Miltenyi | 130-118-960 |
| HLA-B7/B27 | REA176 | FITC | Miltenyi | 130-118-332 |
| HLA-ABC | W6/32 | APC | Biolegend | 311410 |
| Blimp1 | 6D3 | PE-CF594 | BD Biosciences | 565274 |
| Ki67 | Ki-67 | APC | Biolegend | 350513 |
| Granzyme B | QA16A02 | AF700 | Biolegend | 372222 |
| Granzyme K | GM26E7 | PE/Cy7 | Biolegend | 370515 |
| CD107a | H4A3 | BV605 | Biolegend | 328634 |
| Perforin | B-D48 | APC | Biolegend | 353312 |
| KLRC1 | S19004C | AF647 | Biolegend | 375105 |
| CD19 | HIB19 | PCP-Cy5.5 | Biolegend | 302230 |
| CD20 | 2H7 | PCP-Cy5.5 | Biolegend | 302326 |
| CD56 | HCD56 | PCP-Cy5.5 | Biolegend | 218321 |
| CD14 | 63D3 | PCP-Cy5.5 | Biolegend | 367110 |
| CD4 | RPA-T4 | APC/Cy7 | Biolegend | 300518 |
| CD137 | 4B4-1 | BV605 | Biolegend | 309822 |
| OX40 | Ber-ACT35 | PE/Cy7 | Biolegend | 350012 |
| CD25 | BC96 | PE/Cy5 | Biolegend | 302607 |
| CD45 | HI30 | BUV395 | BD Biosciences | 563792 |
| CFSE | NA | NA | Biolegend | 423801 |
| eFluor^TM^ 450 | NA | NA | ThermoFisher | 65-0842-85 |
| Cell Trace Violet^TM^ | NA | NA | ThermoFisher | C34571 |
| **Immunofluorescence imaging** | | | | |
| **Antibody** | **Clone** | **Fluorochrome** | **Supplier** | **Catalog #** |
| HLA-B7 | EPR2623(2) | None | Abcam | ab247757 |
| CD3 | 3F3A1 | AF594 | Invitrogen^TM^ | CL594-60181 |
| Granzyme B | 23H8L20 | None | Invitrogen^TM^ | PI701395 |
| CRIP1 |  | None | SigmaAldrich | HPA042462-25UL |
| Anti-mouse |  | AF647 | Cell Signaling Tech. |  |
